# Using routine emergency department data for syndromic surveillance of acute respiratory illness in Germany, week 10-2017 to 10-2021

**DOI:** 10.1101/2021.08.19.21262303

**Authors:** T. Sonia Boender, Wei Cai, Madlen Schranz, Theresa Kocher, Birte Wagner, Alexander Ullrich, Silke Buda, Rebecca Zöllner, Felix Greiner, Michaela Diercke, Linus Grabenhenrich

**Affiliations:** Robert Koch Institute, Department for Infectious Disease Epidemiology, Berlin, Germany; Charité – Universitätsmedizin Berlin, corporate member of Freie Universität Berlin and Humboldt Universität zu Berlin, Institute of Public Health, Berlin, Germany; Robert Koch Institute, Department for Methodology and Research Infrastructure, Berlin, Germany; Health Protection Authority, Frankfurt am Main, Germany; Department of Trauma Surgery, Otto von Guericke University Magdeburg, Magdeburg, Germany; AKTIN – Emergency Department Data Registry, Magdeburg/Aachen, Germany; Institute for Occupational and Maritime Medicine (ZfAM), University Medical Center Hamburg-Eppendorf (UKE), Germany

**Keywords:** Public Health Surveillance, COVID-19, Respiratory Tract Infections, Emergency Service, Hospital

## Abstract

**Background:** The Coronavirus disease 2019 (COVID-19) pandemic expanded the need for timely information on acute respiratory illness on the population level.

**Aim:** We explored the potential of routine emergency department data for syndromic surveillance of acute respiratory illness in Germany.

**Methods:** We included routine attendance data from emergency departments who continuously transferred data between week 10-2017 and 10-2021, with ICD-10 codes available for >75% of the attendances. Case definitions for acute respiratory illness (ARI), severe ARI (SARI), influenza-like illness (ILI), respiratory syncytial virus disease (RSV) and COVID-19 were based on a combination of ICD-10 codes, and/or chief complaints, sometimes combined with information on hospitalisation and age.

**Results:** We included 1,372,958 attendances from eight emergency departments. The number of attendances dropped in March 2020, increased during summer, and declined again during the resurge of COVID-19 cases in autumn and winter of 2020/2021. A pattern of seasonality of acute respiratory infections could be observed. By using different case definitions (i.e. for ARI, SARI, ILI, RSV) both the annual influenza seasons in the years 2017-2020 and the dynamics of the COVID-19 pandemic in 2020-2021 were apparent. The absence of the 2020/2021 flu season was visible, parallel to the resurge of COVID-19 cases. The percentage SARI among ARI cases peaked in April-May 2020 (17%) and November 2020-January 2021 (14%).

**Conclusion:** Syndromic surveillance using routine emergency department data has the potential to monitor the trends, timing, duration, magnitude and severity of illness caused by respiratory viruses, including both influenza and SARS-CoV-2.

## Introduction

The response to the Coronavirus disease 2019 (COVID-19) pandemic expanded the need for timely information on acute respiratory illness on the population level, to trigger public health action, and support healthcare planning. Therefore, robust surveillance systems of respiratory illnesses are essential to monitor the burden of disease, and therewith inform public health to take appropriate and timely measures. The effects of the COVID-19 pandemic, as well as the associated public health and social measures to contain the spread of the virus, have affected the transmission of not only seasonal influenza, but also other respiratory viruses in Germany (1, 2). These changes in the epidemiological situation affect routine surveillance systems for monitoring the epidemiology and virology of respiratory illnesses (3). Therefore, further investigation of the impact on population health, as well as the healthcare system is required.

As public health institutes are planning to transition from COVID-19 emergency surveillance to routine surveillance of all respiratory pathogens, syndromic surveillance of acute respiratory illness – i.e. cross-pathogen symptomatic cases – plays an important role (4). In Germany, certain syndromic surveillance systems for acute respiratory illnesses (ARI) have been implemented before the pandemic: internet-based syndromic monitoring of ARI in the general population (*GrippeWeb*, “FluWeb”)(5); sentinel surveillance of ARI consultations in primary care, including virological surveillance (Influenza Working Group(6)) and sentinel surveillance of severe acute respiratory illnesses (SARI) among hospitalised patients using ICD-10 diagnostic codes (ICOSARI)(7). In addition, defined respiratory diseases as COVID-19 and influenza are monitored through mandatory notifications by clinicians and laboratories, within the framework of the Protection against Infection Act.

The availability of electronically collected routine medical data, such as emergency department data, allows for an extended approach in applied epidemiology and public health surveillance (8). Syndromic surveillance using routine emergency department data can be a fast approach to report and detect changes in healthcare utilization, without creating an additional administrative burden. In the United Kingdom (9), France (10) and other countries, syndromic surveillance systems based on emergency department data have been established (11, 12), and have previously provided timely public health insight at extraordinary times such as extreme weather events (13) or mass gatherings (e.g. the Olympic Games in 2012 (14)), as well as during the COVID-19 pandemic (15).

Previous analyses of emergency department data in Germany successfully detected syndromes to monitor unspecific gastrointestinal infections, including their seasonal fluctuation (16), as well as a first description of ARI cases (17). Since end of June 2020, the weekly Emergency Department Situation Report presents data from the routine documentation of up to 21 selected emergency departments in Germany (18). These reports showed that in parallel to the COVID-19 pandemic and the associated public health and social measures starting mid-March 2020, attendances dropped(19). Nation-wide public health and social measures commenced in February-March 2020, including a “contact ban” allowing a maximum of two people, closure of most public spaces including schools, non-essential shops, and many business (20-22). By April-May 2020, physical distancing measures were gradually eased. During the summer months of 2020, emergency department attendances partially increased again (19). Large-scale testing focusing on symptomatic suspected cases, contact persons and vulnerable settings stayed in place, as well as the use of face-masks, physical distancing and a quarantine for people retuning from areas classified as high-risk. During the resurge of cases in the fall of 2020, many public health and social measures were re-implemented, including closure of most public spaces (including non-essential shops and hospitality), a working from home policy, and the call to reduce personal contacts to the bare minimum (21). By December 2020, non-essential shops and schools and were also closed. During the resurge of COVID-19 cases and implementation of public health and social measures during the fall and winter of 2020-2021, the number of attendances decreased again to about one quarter below the 2019 average Gradual lifting of measures started from March 2021, followed by the introducing a “3G” policy for shops, sports, and hospitality (i.e. requiring proof of either a negative COVID-19 lateral flow test (*‘Getestet’*), completed course of vaccination (*‘Geimpft’*), or recent recovery (*‘Genesen’*) (21). Emergency attendances remained below 2019 average until the time of analysis and writing (April 2021) (18).

In this work, we aimed to describe emergency department attendances for acute respiratory illness in Germany over time, for the purpose of the development and implementation of syndromic surveillance. Findings of this study facilitated the implementation of surveillance indicators of acute respiratory illness in the weekly emergency department surveillance reports (23), and cross-pathogen syndromic surveillance of symptomatic cases of respiratory illness in Germany overall(4). Furthermore, we explore the impact of the COVID-19 pandemic, and its associated public health and social measures, on these presentations of acute respiratory illness.

## Methods

### Study design

We conducted a retrospective observational study to explore the potential of routine emergency department data for syndromic surveillance of acute respiratory illness in Germany. Following a process of data curation, quality management and exploration, and defining syndromic case definitions, we described emergency department attendances over time for the period of 6 March 2017 up to and including 13 March 2021, to provide insight into presentation for acute respiratory illness.

### Setting

Data used in this analysis was provided through two networks of German emergency departments which continuously transfer anonymized routine health data to the Robert Koch Institute (RKI, Germany’s national public health institute)(24): the AKTIN – Emergency Department Data Registry (25) and the ESEG-project (*“Erkennung und Steurung Epidemischer Gefahrenlagen”*; translation: detection and control of epidemic situations http://www.rki.de/eseg). Germany has over 1,000 emergency departments that differ in size and level of care (basic/extended/comprehensive emergency care) (26). There are no barriers to accessibility, as patients can be admitted by ambulance, referred by a general practitioner or even by themselves as “walk-in” patients. Emergency departments participate and share data on a voluntary basis. At the RKI, these routine health data from the different sources are merged and processed using the NoKeDa (*NotaufnahmeKernDatensatz*) data model (27), when the date of visit, age, and sex were available. Observations were recorded by visit (attendance) and not by person; multiple visits by the same person cannot be linked.

### Study population

Emergency attendance data was exported from the RKI server on 13 April 2021, using SUMO-DB Release 3.0.3 (database population). In addition to the database release, data quality summary reports were generated, including information on completeness of attendances and variables. The data availability and quality could be assessed, based on the descriptive statistics and data visualisation of data quality summary. For the purpose of this work, a subset of emergency departments was included (study population): departments who continuously transferred attendance data (at least one admission per day) for the period of 6 March 2017 (week 10-2017) up to and including 13 March 2021 (week 10-2021), with ICD-10 diagnostic codes for >75% of the attendances.

### Data protection and ethics

In AKTIN and ESEG, individual patient consent is not provided for the context of the emergency situation and the technical and organizational measures provided. The AKTIN-register received medical ethical approval from the Ethics Committee of the Otto von Guericke University Magdeburg, Medical Faculty (160/15) (28). The AKTIN scientific board approved regular/daily data query and transmission to RKI (Project-ID 2019-003). Due to the anonymized nature of the data, an ethical approval was not necessary for ESEG data, as disclosed by the ethics committee of the physician’s chamber Hesse. The data were transferred anonymously. The data model uses a minimal set of data points, or categorisation of specific data elements (e.g. age in years and groups, instead of date of birth), which ensures personal data protection, and defines an approved list of variables. The data protection officer of the RKI has approved the use of anonymized case-based emergency department data using the NoKeDa model for research purposes (BDS/ISB, 09-01-2019).

### Variables

The following variables were included for analysis: emergency department/hospital, age (collected in age-groups: 0-2 years, 3-4 years, followed by 5-year groups, and 80+ years), sex (male/female/other), triage (codes 1 to 5, higher code meaning less urgent), diagnosis (ICD-10 codes, when labelled confirmed/suspected), chief complaints (according to the Canadian Emergency Department Information System – Presenting Complaint List [CEDIS-PCL](29) or Manchester Triage System [MTS] (30)), referral, mode of transport, disposition (e.g. hospital admission) and isolation. Potential missing data refers to the absence of documentation; all data are based on routine emergency care at the respective sites, based on routine documentation practices.

### Case definitions

We defined a set of case definitions in agreement with the established syndromic surveillance systems of acute respiratory infections at RKI and based on (inter)national standards (31, 32). Case definitions were based on a combination of ICD-10 diagnostic codes, or chief complaints, sometimes combined with disposition and age. Case criteria and case classifications (i.e. probable, confirmed) for acute respiratory illness (ARI), severe acute respiratory illness (SARI), influenza-like illness (ILI), respiratory syncytial virus (RSV) and Coronavirus disease 2019 (COVID-19) are summarised in **Supplemental Table 1**. Combined case definitions were used for the primary analysis; i.e. cases of ILI, RSV and COVID-19 were either probable and/or confirmed.

### Statistical analysis

Attendances and cases were analysed as aggregated counts by week. Weekly numbers of all cases that met the case definitions (see above and **Supplemental Table 1**) were plotted as absolute counts, as well as number of cases per 1,000 attendances to adjust for fluctuations in attendances (i.e. a change in the denominator) over time. Cases of ARI and SARI were explored as individual indicators, as well as through calculating the percentage of severe cases among ARI cases (SARI/ARI). Probable and confirmed cases of ILI, RSV-Disease and COVID-19 were explored as separate and combined overall case numbers (probable + confirmed cases).

The number of weekly attendances were described and summarised for the period before and during the COVID-19 pandemic (2 March 2020, week 10/2020). These different phases of the COVID-19 pandemic in Germany have been categorised following Schilling et. al (33): 0) pre-pandemic and sporadic cases up to and including week 09/2020, 1) first COVID-19 wave week 10/2020-20-2020; 2) summer plateau week 21/2020-39/2020; 3) second COVID-19 wave week 40/2020-10/2021. Because the number of weekly attendances were not normally distributed, Wilcoxon rank sum test was used for simple comparisons of the distribution of weekly cases per 1,000 attendances before and during the pandemic. Next, time series were decomposed to visually investigate the seasonality and trend over time; a decomposition of the COVID-19 time series of one year was not possible because the series was too short (less than two periods).

Data were analysed using R statistical software, using the *tidyverse, xts* and *zoo* packages (34). The aggregated data of the attendances and cases (ARI, SARI, ILI, RSV, COVID-19) by week used for the analysis, are provided in the **Supplemental Material**.

## Results

Consequently, the study population included data of 1,372,958 attendances between 6 March 2017 (week 10-2017) up to and including 13 March 2021 (week 10-2021) from eight emergency departments (**Table 1**). The included departments provided comprehensive (5 departments) and extended (3 departments) emergency care in seven federal states of Germany (Baden-Württemberg, Bavaria, Berlin, Brandenburg, Hesse, Lower Saxony, Saxony).

**Table 1.**
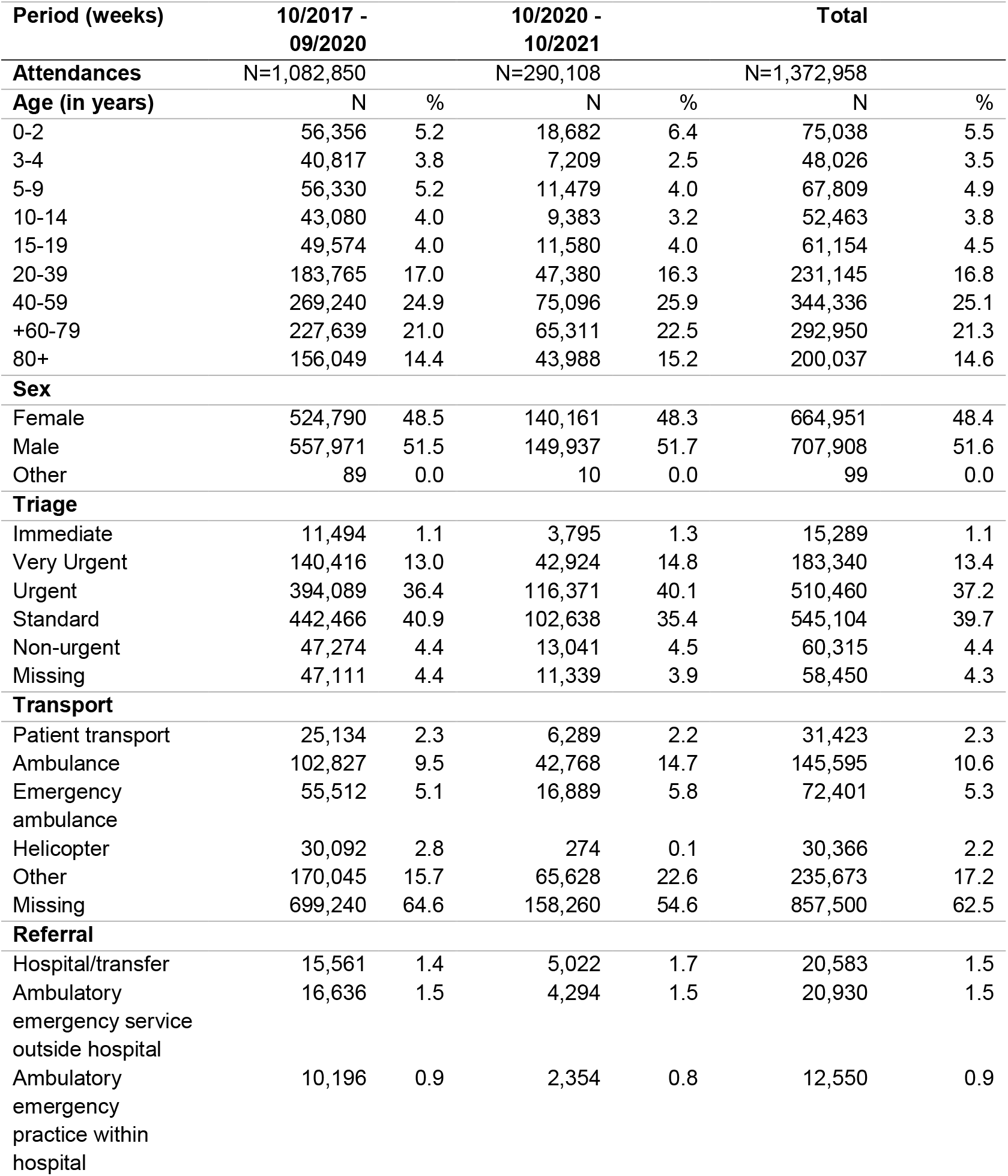
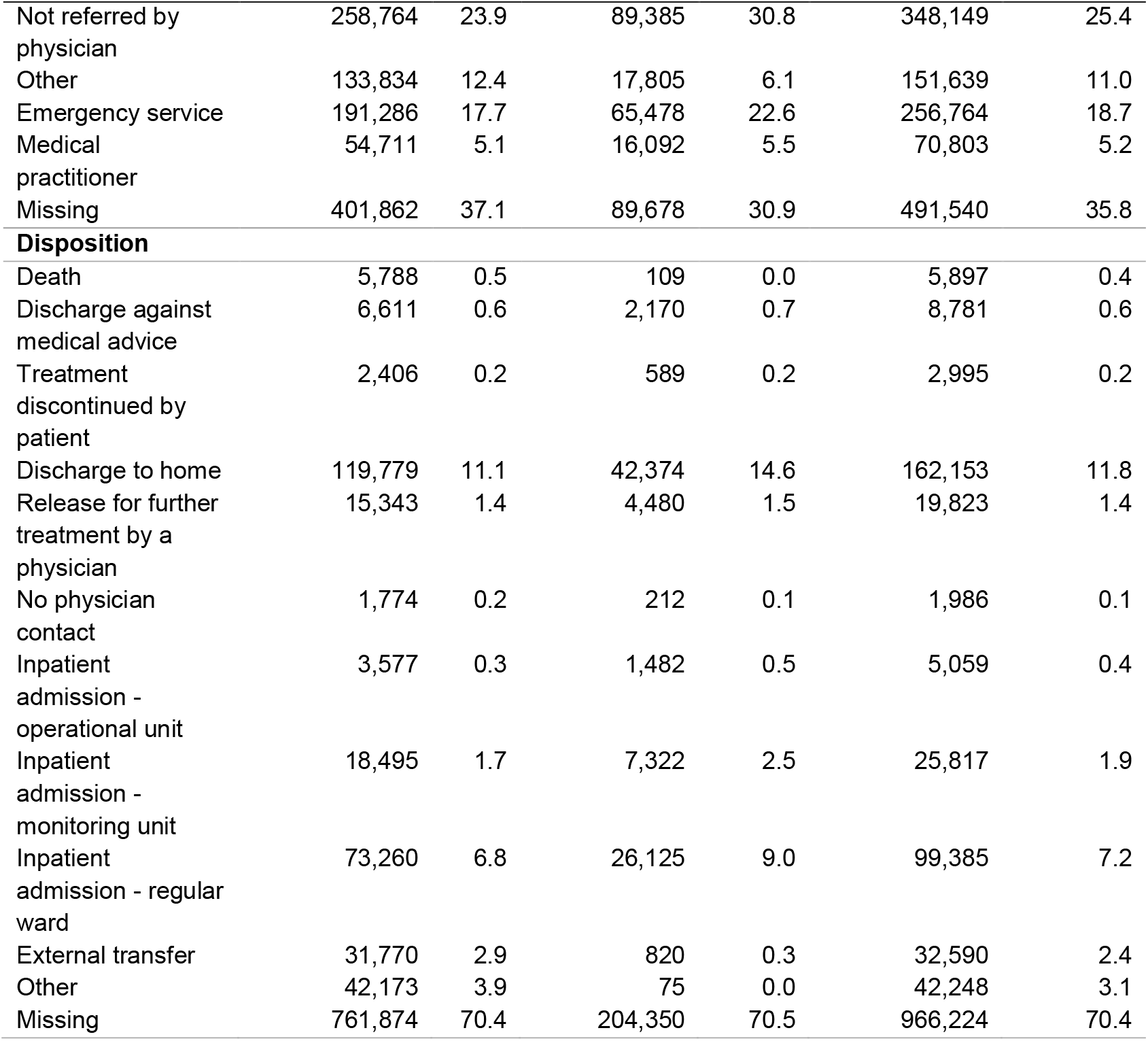
**Summary of total emergency department attendances, between 6 March 2017 (week 10-2017) up to and including 13 March 2021 (week 10-2021), for the full observation period, and stratified for the time before and during the COVID-19 pandemic**.

The general characteristics of the emergency department attendances are summarized in Table 1 (**Table 1**). The overall sex (52% female) and age (6% under 2 years, 8% 3-9 years; 8% 10-19 years; 17% 20-39 years; 25% 40-59 years; 21% 60-79 years; 15% 80+ years) distributions remained similar over the full observation period, with the largest decrease in attendances under those 0-19 years of age (**Supplemental Table 2 and Supplemental Figure 1**).

Over time, the absolute number of weekly emergency department attendances dropped during the first and second wave of the COVID-19 pandemic in Germany (**Figure 1**). The median number of attendances per week dropped from 6,886 (interquartile range [IQR]: 6,733 - 7,104) before the pandemic (<week 10/2020), to 4,741 (IQR: 4,498 – 5,124; *P* <.001), during the first COVID-19 wave. Following a recovery phase approaching pre-pandemic attendance rates (median 6,133; IQR: 6,010 - 6,202; *P* <.001) during the summer plateau (week 21/2020-39/2020), the weekly attendances declined again to 4,950 (IQR: 4,512-5,212; *P* <.001) during the second COVID-19 wave during autumn and winter of 2020/2021 (week 40/2020-10/2021). The weekly attendances differed when comparing the respective phases to the pre-pandemic period, as summarised in **Supplemental Figure 2**. The highest number of 8,412 attendances per week was counted in week 09/2018, and the lowest number of 4,096 attendances per week was counted in week 12/2020.

**Figure 1.**
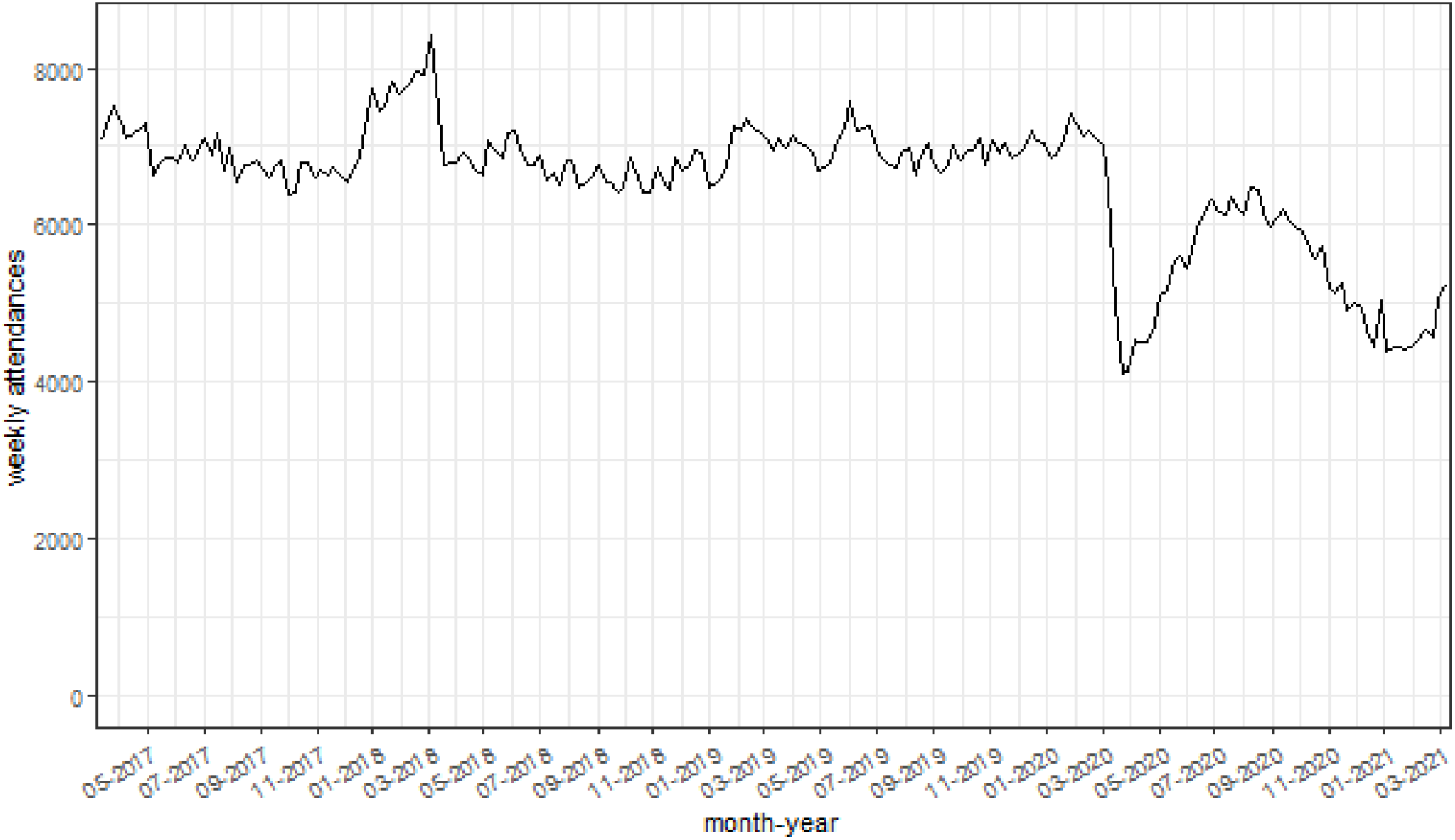
Weekly emergency department attendances, between 6 March 2017 (week 10-2017) up to and including 13 March 2021 (week 10-2021).

### Cases of ARI, SARI, ILI, RSV-disease and COVID-19 over time

During the full, four-year observation period, seasonal patterns were visible for ARI, SARI, ILI, and RSV-disease cases per week, per 1,000 attendees (**Figure 2**), as well as per absolute weekly case counts (**Supplemental Figure 2**). The decomposed time series analysis suggested seasonal factors of ARI, SARI, ILI, and RSV cases (**Figure 3**).

**Figure 2.**
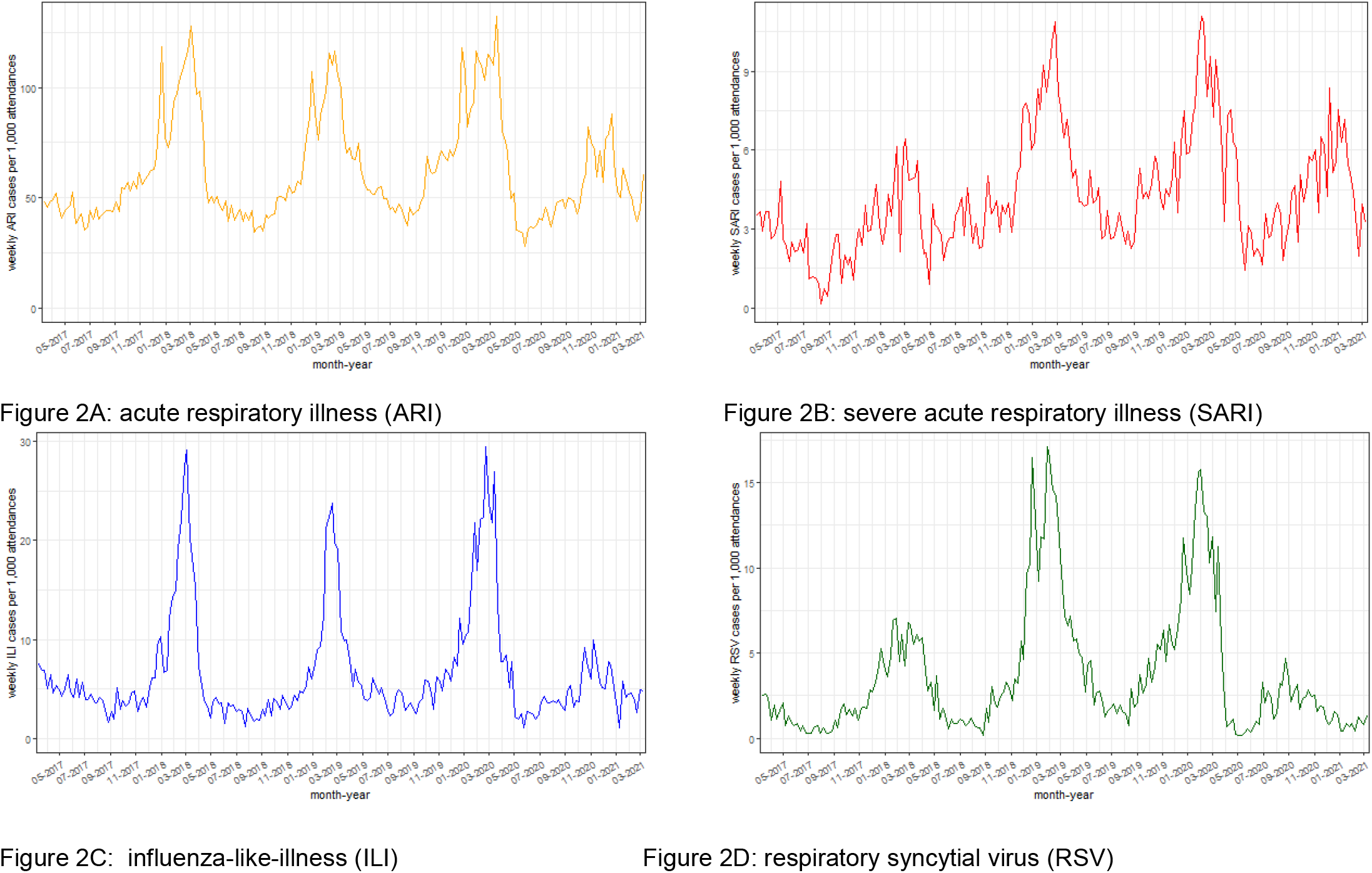
Weekly case counts (ARI, SARI, ILI, RSV) per 1,000 emergency department attendances, between 6 March 2017 and 13 March 2021.

**Figure 3.**
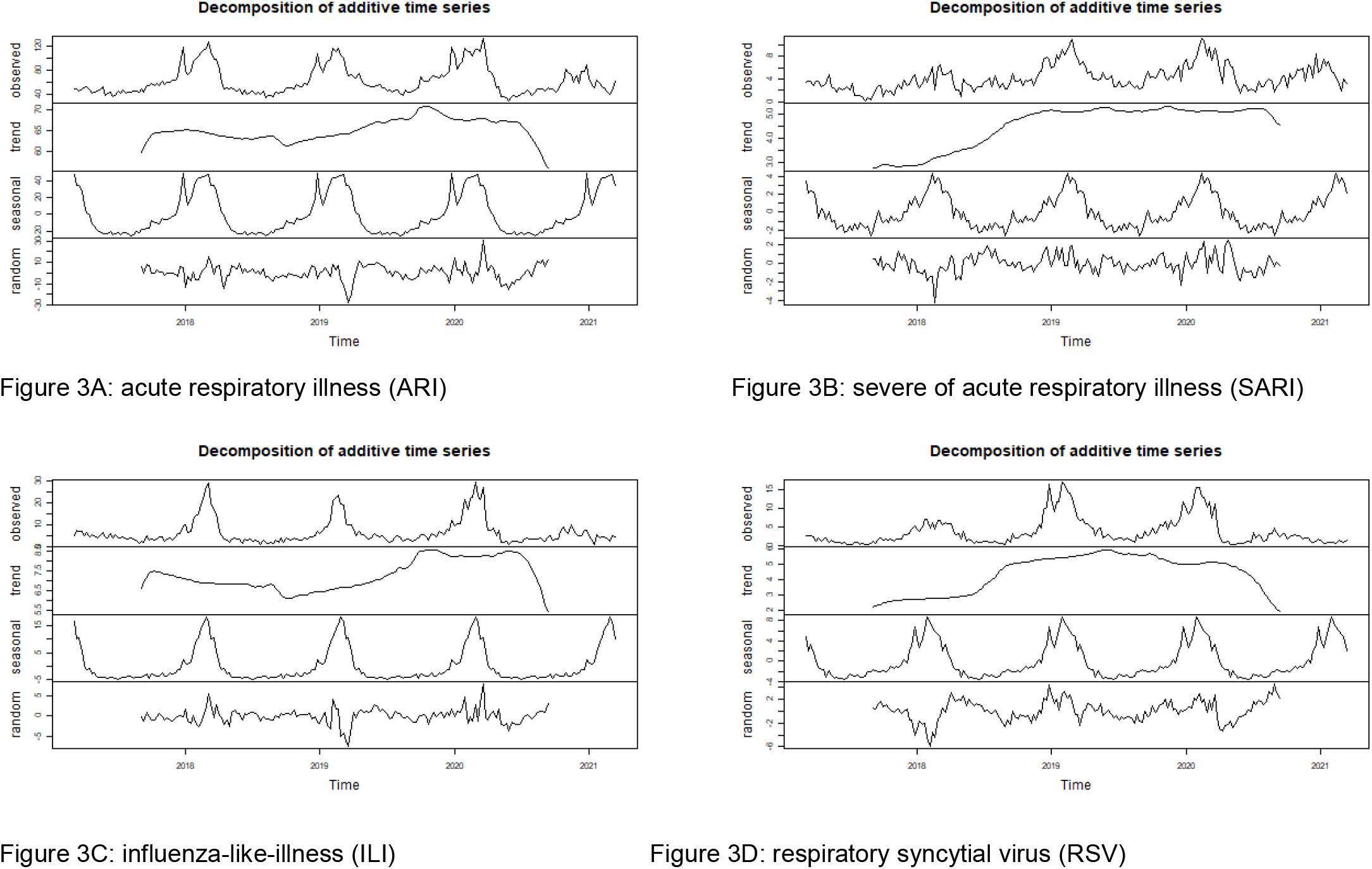
Decomposed time series of the weekly case counts, per 1,000 emergency department attendances, between 6 March 2017 and 13 March 2021.

In the winter of 2020/2021, both the absolute and relative ARI case counts were lower than in previous years, and showed a downward trend (**Figure 2a and 3a**). Conversely, the trend of SARI cases remained fairly stable over time, with a small downward trend in the final months of winter (**Figure 2b and 3b**). This pattern was also visible when assessing the % of severe (i.e. SARI) among ARI cases (**Figure 4**). The % of SARI cases among ARI cases fluctuated around a median of 6.9% (IQR: 5.3-8.3), was lower in the year 2017 (median 4.7% [IQR: 3.4-6.2]), fairly stable in the years 2018 (6.6% [IQR: 5.1-8.0]) and 2019 (7.4% [IQR: 6.4-8.4]). The %SARI cases among ARI cases occasionally exceeded 10%, but was only observed in single weeks during pre-pandemic times: week 18/2017, week 30/2018, week 37/2018; week 8/2019, week 20/2019, and week 37/2019. Twice during the pandemic, the % of SARI cases among ARI cases exceeded 10% for five subsequent weeks in a row: week 15/2020 to 19/2020 with a peak at week 18/2020 (17.1%) and week 48/2020 to week 2/2021 with a peak at week 53/2020 (14.2%), as displayed in **Figure 4**.

**Figure 4.**
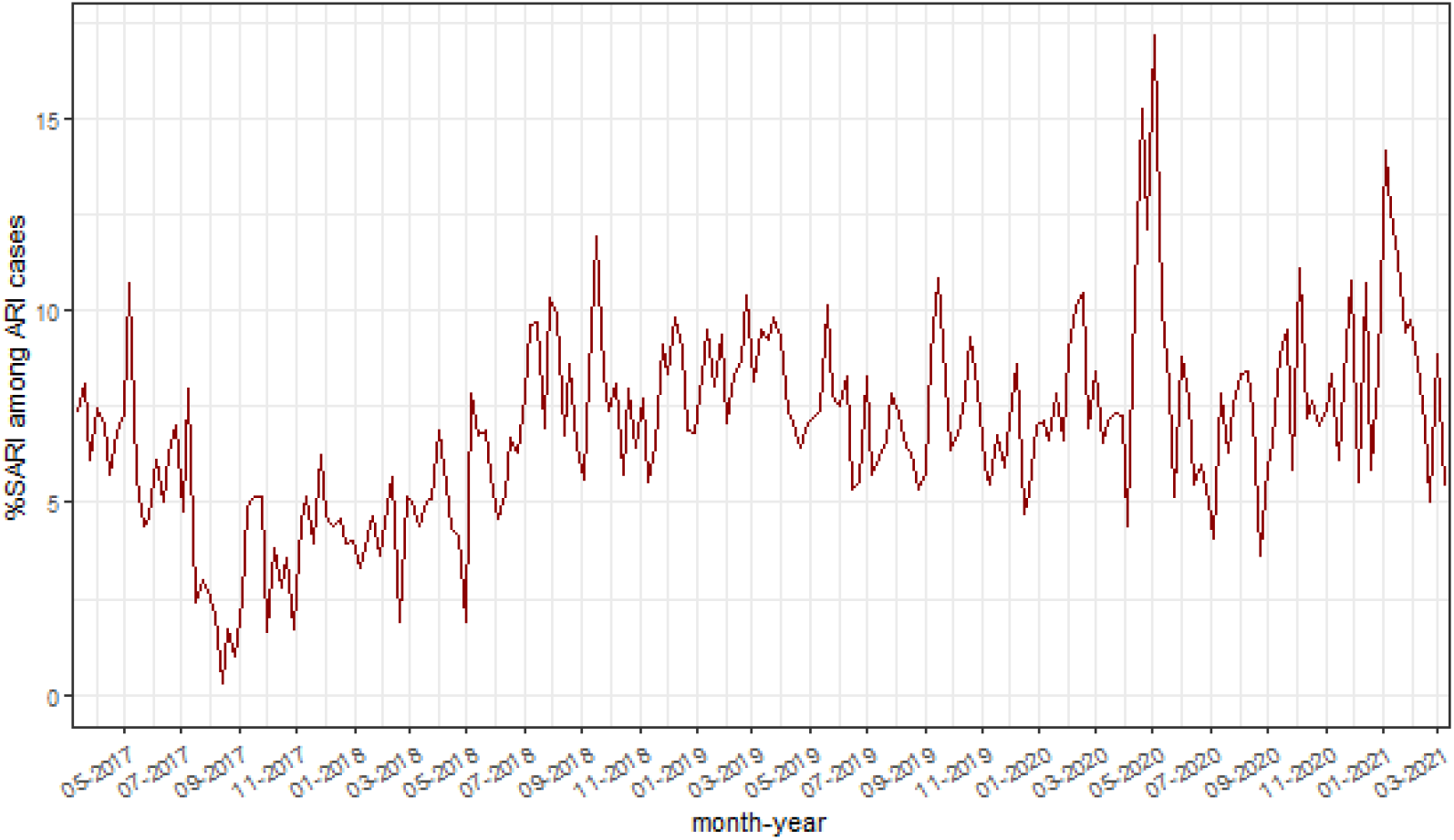
Proportion (%) of severe acute respiratory illness (SARI) cases among cases of acute respiratory illness (ARI), between 6 March 2017 and 13 March 2021.

After March 2020, the expected seasonal increases in cases of ILI and RSV cases was reduced (**Figure 2c-d and 3c-d**). In parallel, COVID-19 cases were recorded from March 2020 onwards (**Figure 5**). Visual inspection of COVID-19 cases showed two periods of increased case numbers, despite the drop of overall attendances, during April-May of 2020 and a resurge of cases starting in October 2020, which was decreased again at the end of the observation period (March 2021). Overall, 17% of COVID-19 cases were also cases of SARI (all COVID-19 cases are by definition ARI cases). Of note, weekly ILI, RSV and COVID-19 cases are plotted by probable and possible case classifications, as well as the combined case definition, in **Supplemental Figure 4**.

**Figure 5.**
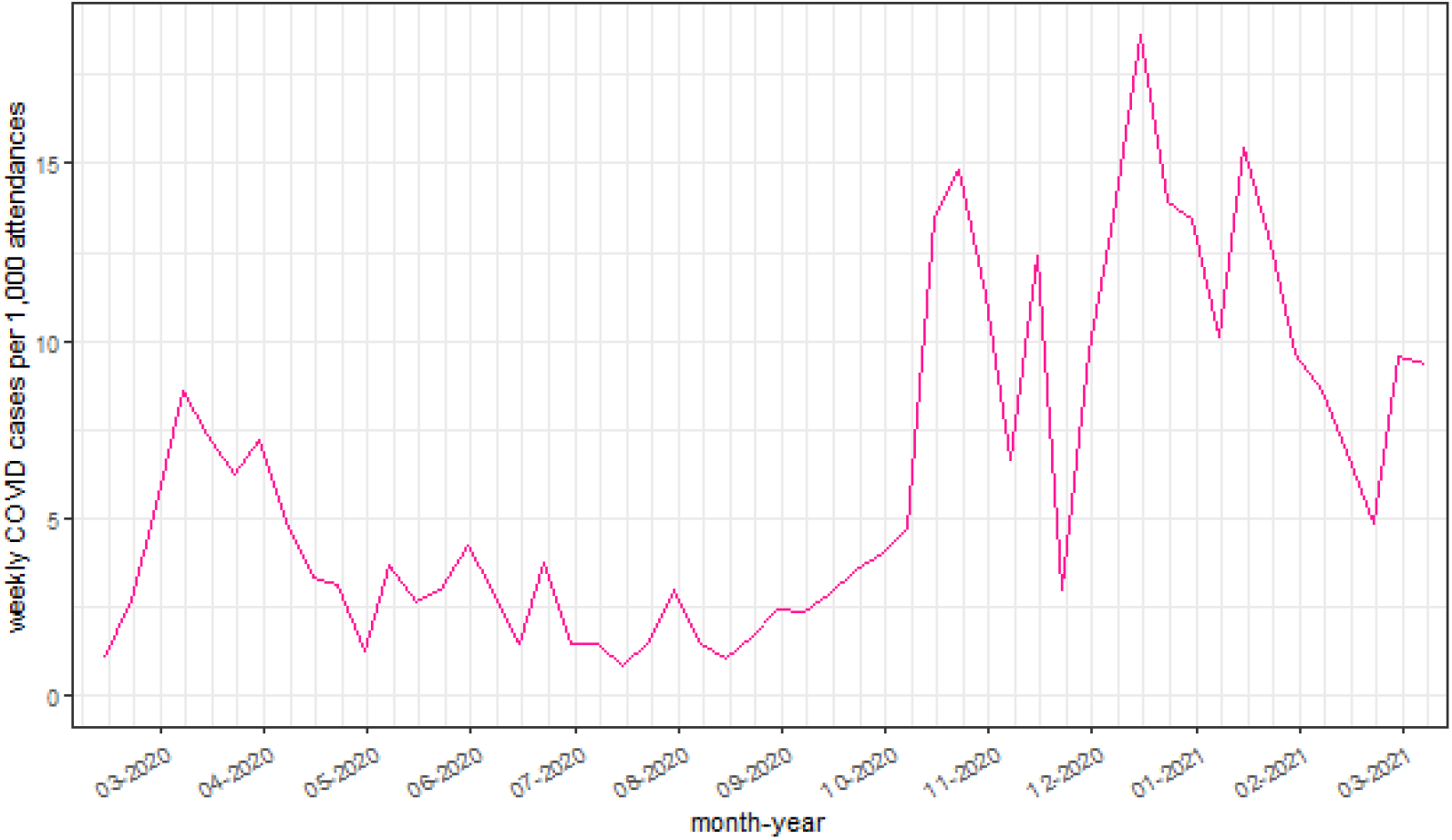
Summary of COVID-19 cases per week, per 1,000 emergency department attendances, between 6 March 2020 and 13 March 2021.

## Discussion

This descriptive study indicates proof of principle for syndromic surveillance of acute respiratory illness in emergency departments in Germany, directly feeding into routine emergency department surveillance reporting (23). Based on a selection of eight departments by voluntary participation, a clear pattern of seasonality respiratory illness (i.e. ARI, SARI, ILI and RSV-disease) could be observed. In addition, both the annual influenza seasons in the years 2017-2020 and the dynamics of the COVID-19 pandemic in 2020-2021 were apparent. The more severe flu 2017/2018 season was visible in the emergency department data, through a higher peak of attendances due to ARI compared to the 2018/2019 and 2019/2020, which is consistent with the severity of the three influenza seasons observed in the other syndromic surveillance systems (33, 35, 36). Of note, the abrupt and early end of the 2019/2020 flu wave could be visually identified, as described based on sentinel surveillance in Germany (37), as well as in Denmark, Norway and Sweden (38). The striking absence of a flu season in 2020/2021 (1, 39), was also visible, in parallel to the resurge of COVID-19 cases during the fall and winter in 2020/2021. In 2020 and the beginning of 2021, the percentage of SARI cases among ARI cases peaked twice for five subsequent weeks in April-May 2020 and November 2020-January 2021, which was the same time the first wave and resurge of COVID-19 cases peaked in Germany (40). The reduction in ILI and RSV cases during the pandemic period could, at least in part, be explained by the influence of public health and social measures on overall disease transmission, as observed by other surveillance systems in Germany (1, 2), and internationally (41).

Overall emergency department attendances dropped substantially following the start of the COVID-19 pandemic in March 2020, increased during summer, and declined again during the resurge of the pandemic during the autumn and winter of 2020/2021. The shift in attendances required a baseline adjustment for case reporting (change in denominator), by standardising the case count per 1,000 attendances. This reduction in attendances could have been partially due to the advisories to reduce physical contact with healthcare services, and to use telephone services instead (at least before visiting), when possible. This observation fits the overall reduction in hospitalisation as reported by other German hospitals (42, 43). In addition, a reluctance to visit health care facilities due to a fear to be exposed to SARS-CoV-2 (44), could also be a reason for a reduction in emergency department attendances. Similar trends were observed in emergency departments in other countries, including the UK (45) and USA (46).

The findings of this study should be interpreted taking the following limitations into account. First, the selected data are a convenience sample and are not representative for emergency departments in Germany as a whole. Differences on the level of individual departments, as well as differences between emergency departments, played a limited role in the analysis and interpretation. This study aimed to explore the possibility and feasibility to use these routine data for public health surveillance. In addition, structural changes at the department, including active interreference with patient flows with respiratory complaints, could have affected reported attendances and case counts. To fully understand trends in the data, being it due to a change in infection dynamics, healthcare seeking behaviour, the COVID-19 pandemic and public health and social measures, or structural changes at the emergency department, more information is needed. This type of investigation is, however, beyond the scope of this work. The strength of the current approach of passive surveillance using routine documentation has its own advantages being fast (timely reporting) without creating an additional burden on the healthcare workers. Furthermore, case definitions were based on routine documentation, designed for triage and clinical documentation in the emergency department, not designed for surveillance purposes. The syndromic case definitions heavily rely on clinical presentation, and laboratory diagnosis is often not available, or not foreseen. Shortness of breath, for example, may be a cardiac symptom as well, and is not always caused by a respiratory pathogen (47). Furthermore, information on hospitalisation (disposition), a key variable for SARI cases, was not always available, leading to potential underreporting of SARI cases. In addition, ICD-10 codes for COVID-19 diagnosis were not available from the beginning of the pandemic, leading to underreporting of the true number of cases.

As of November 2021, the syndromic surveillance indicators based on the ARI, SARI and confirmed-ILI case definitions in this paper are included in the weekly reporting of the Emergency Department Surveillance in Germany, by the RKI (23). This updated version of the report includes a data quality table of each variable of the current week. Moving forward, the current ongoing, systematic collection, analysis, interpretation, and dissemination of emergency department data for use in public health action, needs to be evaluated, to make targeted recommendations to improve quality, efficiency, and usefulness of the system (48). Especially in the context of the COVID-19 pandemic, targeted analyses looking into the potential shifts in attendances during different phases of the pandemic are ongoing. Triangulation of data sources and formal comparison with established surveillance systems, including COVID-19 case notifications (49), and other syndromic and virological surveillance systems for acute respiratory infections (5, 7), will help to address under-ascertainment, to understand changes in consultation behaviour during public health emergencies and improve interpretation of the data.

The COVID-19 pandemic poses a new challenge for syndromic surveillance, adding an additional respiratory pathogen to the set of illnesses under monitoring and surveillance. Syndromic surveillance in emergency departments can help provide oversight of the healthcare utilisation, independent of the pathogen, for overall public health surveillance and healthcare monitoring (50). Moving forward, increased COVID-19 vaccination roll-out and subsequent (de-escalation of) public health and social measures, will have effects on circulation of other respiratory viruses as well. In the near future, this type of almost real-time syndromic surveillance can help to timely inform public health decision making, moving forward in the next phases of the pandemic.

### Conclusions

The current analysis of four years of emergency department data identified syndromic cases of (severe) acute respiratory illness, and could differentiate between ILI, RSV and COVID-19 cases, which should be further investigated. Clear patterns of seasonality and trends over time were identified, for all case definitions, in line with findings reported from established ARI, SARI, ILI and virological surveillance systems (51). Syndromic surveillance of ARI and SARI in the emergency department could contribute to the overall burden estimates of acute respiratory infections in the upper and lower respiratory tract, which is essential in the transition transfer from COVID-19 emergency surveillance to routine surveillance of all respiratory pathogens (4). Syndromic surveillance using routine emergency department data has the potential to complement established indicator based, syndromic and virological surveillance systems used to monitor the timing, duration, magnitude and severity of epidemics caused by respiratory viruses, including RSV, influenza virus and SARS-CoV-2.

## Data Availability

The aggregated data of the attendances and cases (ARI, SARI, ILI, RSV, COVID-19) by week used for the analysis, will be provided (as .csv).

## Notes

**Conflict of interest statement:** None declared.

### Competing Interest Statement

The authors have declared no competing interest.

### Funding Statement

The Robert Koch Institute is an Institute within the portfolio of the Federal Ministry of Health. This work was funded by the Innovation Committee of the Federal Joint Committee (Innovationsfonds des Gemeinsamen Bundesausschusses, G-BA) [ESEG project, grant number 01VSF17034]. The implementation and operation of the AKTIN-Emergency Department Data Registry is funded by the Federal Ministry of Education and Research.

### Author Declarations

In AKTIN and ESEG, individual patient consent is not provided for the context of the emergency situation and the technical and organizational measures provided. The AKTIN-register received medical ethical approval from the Ethics Committee of the Otto von Guericke University Magdeburg, Medical Faculty (160/15). The AKTIN scientific board approved regular/daily data query and transmission to RKI (Project-ID 2019-003). Due to the anonymized nature of the data, an ethical approval was not necessary for ESEG data, as disclosed by the ethics committee of the physicians chamber Hesse. The data were transferred anonymously. The data model uses a minimal set of data points, or categorisation of specific data elements (e.g. age in years and groups, instead of date of birth), which ensures personal data protection, and defines an approved list of variables. The data protection officer of the RKI has approved the use of anonymized case-based emergency department data using the NoKeDa model for research purposes (BDS/ISB, 09-01-2019).

### Summary of Updates

Title revision

