## Supplemental Material for "Using routine emergency department data for syndromic surveillance of acute respiratory illness in Germany, week 10-2017 to 10-2021"

**Supplemental Table 1. Case definitions, and case classification (if applicable), used for syndromic surveillance based on routine emergency department data.**

| Case | Definition |
|---|---|
| Acute respiratory illness (ARI) | - ICD-10 diagnostic code (any): J00-J22, J44.0, B34.9, U07.1, U07.2<br>**OR**<br>- Chief complaint(s):<br>o [MTS: breathing problems<br>**AND**<br>(elevated temperature/fever **OR** suspected sepsis **OR** productive cough **OR** shortness of breath **OR** elevated breathing)]<br>**OR**<br>o [MTS: sore throat<br>**AND**<br>(elevated temperature/fever **OR** suspected sepsis **OR** special risk of infection **OR** known or suspected immunosuppression **OR** travel i.e. report on recent stay abroad **OR** special risk of infection **OR** airway at risk **OR** short of breathing **OR** light/medium/strong pain **OR** unresponsive child **OR** rapid onset **OR** shock **OR** hypersalivation **OR** stridor **OR** drowsy i.e. altered level of consciousness)]<br>**OR**<br>o [MTS: asthma<br>**AND**<br>(elevated temperature/fever **OR** suspected sepsis)]<br>**OR**<br>o [MTS: feeling unwell **AND i**nsufficient breathing]<br>**OR**<br>o [CEDIS-PCL (any): 661, 154, 653, 651, 103, 104] |
| Severe acute respiratory illness (SARI) | - ICD-10 diagnostic code (any): J09-J22, U07.1, U07.2<br>**AND**<br>- Disposition:<br>o [Inpatient admission: internal transfer, operational unit, monitoring unit, regular ward<br>**OR**<br>o External transfer] |

*Supplemental Table 1. continued.*

| | |
|---|---|
| Influenza-like illness (ILI) | PROBABLE CASE<br>- ICD-10 diagnostic code (any): J06.-, J12.8, J12.9, J18.-, J22<br>**OR**<br>- Chief complaint(s):<br>o [MTS: breathing problems<br>**AND**<br>(elevated temperature/fever **OR** suspected sepsis **OR** productive cough **OR** shortness of breath **OR** elevated breathing)]<br>**OR**<br>o [MTS: sore throat<br>**AND**<br>(elevated temperature/fever **OR** suspected sepsis **OR** special risk of infection **OR** known or suspected immunosuppression **OR** travel i.e. report on recent stay abroad **OR** special risk of infection **OR** airway at risk **OR** short of breathing **OR** light/medium/strong pain **OR** unresponsive child **OR** rapid onset **OR** shock **OR** hypersalivation **OR** stridor **OR** drowsy i.e. altered level of consciousness)]<br>**OR**<br>o [MTS: asthma<br>**AND**<br>(elevated temperature/fever **OR** suspected sepsis)]<br>**OR**<br>o [MTS: feeling unwell **AND** insufficient breathing]<br>**OR**<br>o [CEDIS-PCL (any): 661, 154, 653, 651, 103, 104]<br>**AND**<br>- Fever:<br>o Temperature ≥38 °C<br>**OR**<br>o Chief complaint:<br>▪ MTS: fever<br>**OR**<br>▪ CEDIS-PCL: 852<br>**OR**<br>o [ICD-10 diagnostic code (any): R50.8, R50.9]<br><br>CONFIRMED CASE<br>- ICD-10 diagnostic code (any): J09, J10.-, J11.- |
| Respiratory syncytial virus (RSV) – | PROBABLE CASE<br>- ICD-10 diagnostic code (any): J12.8, J12.9, J18.-, J20.8, J20.9, J21.8, J21.9, J22<br>**AND**<br>- Age ≤2 years<br><br>CONFIRMED CASE<br>- ICD-10 diagnostic code (any): J12.1, J20.5, J21.0, B97.4 |
| Coronavirus disease 2019 (COVID-19) | PROBABLE CASE<br>- ICD-10 diagnostic code (any): U07.1, U07.2<br><br>CONFIRMED CASE<br>- ICD-10 diagnostic code: U07.1 |

**Supplemental Table 2. Age and sex of emergency attendees over time, broken down by subsequent pandemic phases.**

| | ≤W09/2020 (N=1082850) | W10/2020-W20/2020 (N=55212) | W21/2020-W30/2020 (N=59462) | W31/2020-W39/2020 (N=55697) | ≥W40/2020 (N=119737) | Overall (N=1372958) |
|---|---|---|---|---|---|---|
| **Age (years)** | | | | | | |
| 0-2 | 56356 (5.2%) | 3822 (6.9%) | 3767 (6.3%) | 3482 (6.3%) | 7611 (6.4%) | 75038 (5.5%) |
| 3-4 | 40817 (3.8%) | 1591 (2.9%) | 1483 (2.5%) | 1428 (2.6%) | 2707 (2.3%) | 48026 (3.5%) |
| 5-9 | 56330 (5.2%) | 2239 (4.1%) | 2654 (4.5%) | 2467 (4.4%) | 4119 (3.4%) | 67809 (4.9%) |
| 10-14 | 43080 (4.0%) | 1662 (3.0%) | 1979 (3.3%) | 2072 (3.7%) | 3670 (3.1%) | 52463 (3.8%) |
| 15-19 | 49574 (4.6%) | 2073 (3.8%) | 2301 (3.9%) | 2516 (4.5%) | 4690 (3.9%) | 61154 (4.5%) |
| 20-39 | 183765 (17.0%) | 9008 (16.3%) | 9912 (16.7%) | 9392 (16.9%) | 19068 (15.9%) | 231145 (16.8%) |
| 40-59 | 269240 (24.9%) | 14393 (26.1%) | 15604 (26.2%) | 14160 (25.4%) | 30939 (25.8%) | 344336 (25.1%) |
| 60-79 | 227639 (21.0%) | 12393 (22.4%) | 13276 (22.3%) | 11984 (21.5%) | 27658 (23.1%) | 292950 (21.3%) |
| 80+ | 156049 (14.4%) | 8031 (14.5%) | 8486 (14.3%) | 8196 (14.7%) | 19275 (16.1%) | 200037 (14.6%) |
| **Sex** | | | | | | |
| female | 524790 (48.5%) | 26482 (48.0%) | 28608 (48.1%) | 26792 (48.1%) | 58279 (48.7%) | 664951 (48.4%) |
| male | 557971 (51.5%) | 28728 (52.0%) | 30851 (51.9%) | 28902 (51.9%) | 61456 (51.3%) | 707908 (51.6%) |
| other | 89 (0.0%) | 2 (0.0%) | 3 (0.0%) | 3 (0.0%) | 2 (0.0%) | 99 (0.0%) |

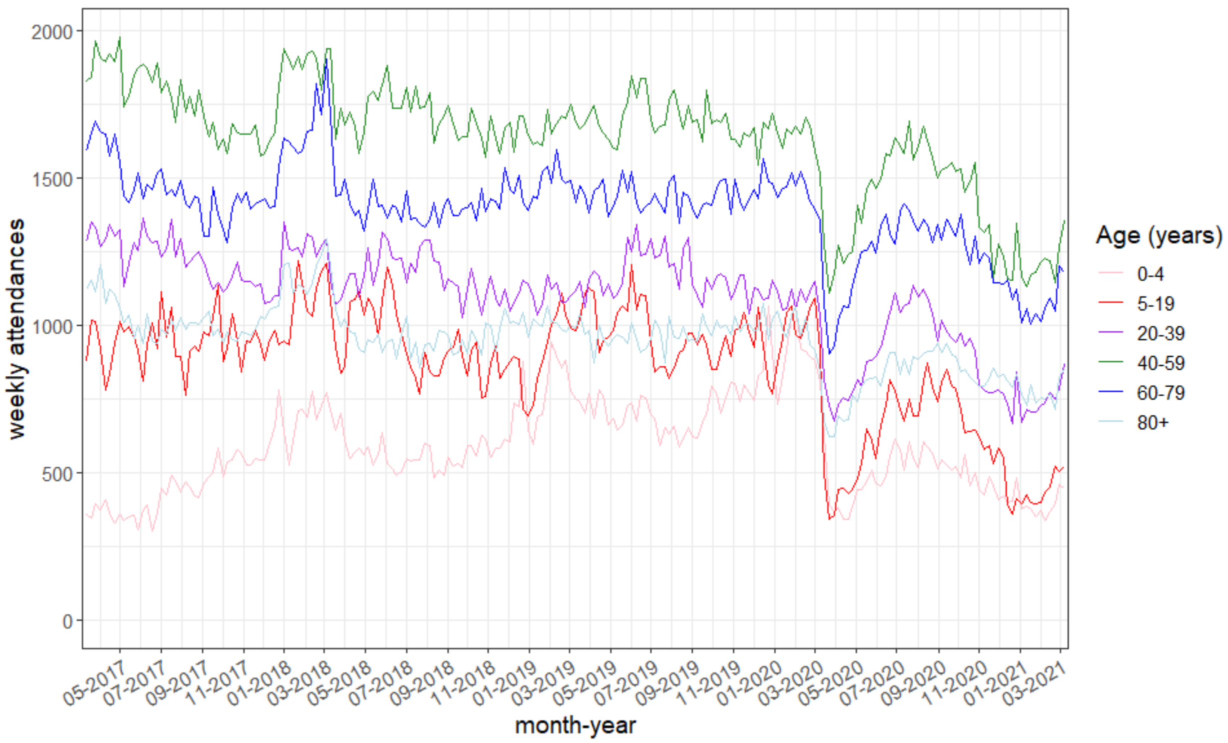


**Supplemental Figure 1. Weekly emergency department attendances, by age group.**

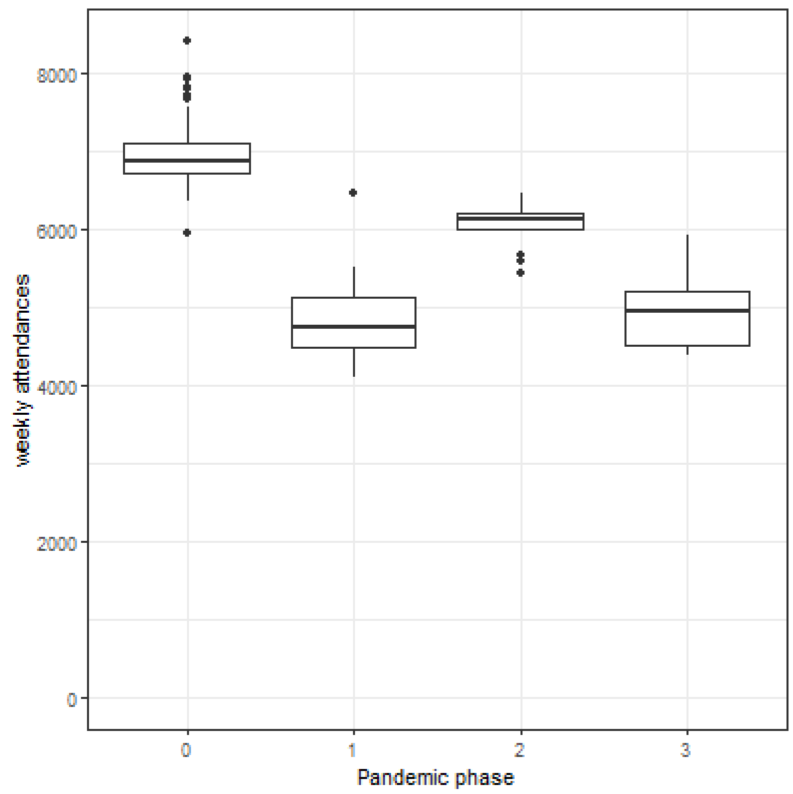


**Supplemental Figure 2. Weekly emergency department attendances, a) before and during the pandemic, broken down by subsequent pandemic phases.**
All comparisons: Wilcoxon rank sum *P* <.001

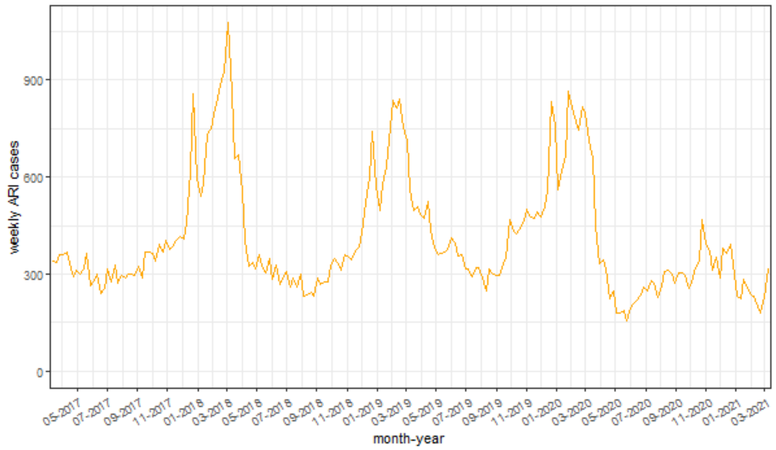


Supplemental Figure 3A

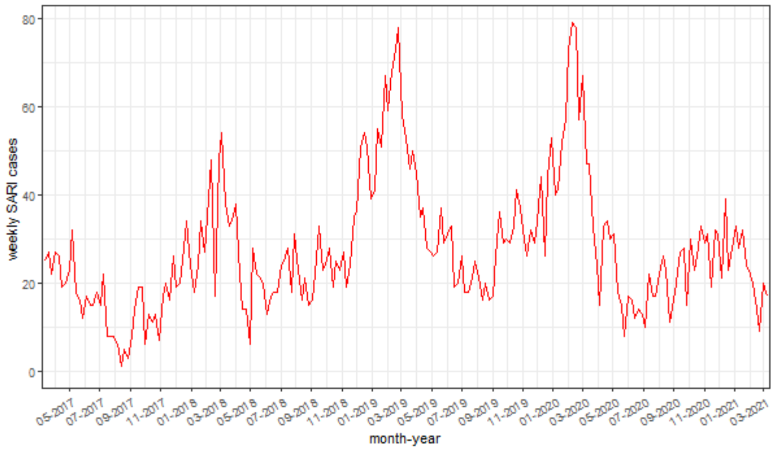


Supplemental Figure 3B

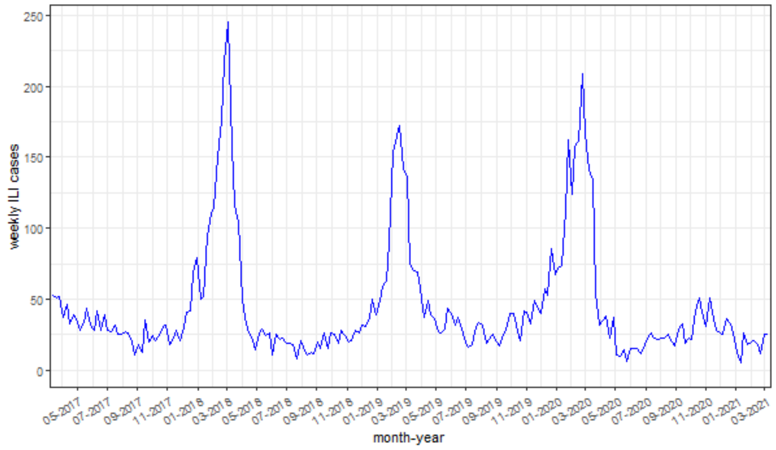


Supplemental Figure 3C

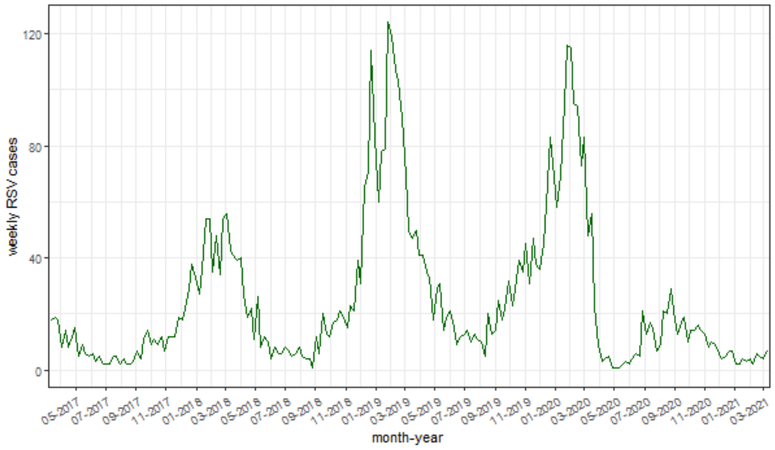


Supplemental Figure 3D

**Supplemental Figure 3. Weekly case count of acute respiratory illness (ARI; 3A), severe acute respiratory illness (SARI; 3B), influenza-like-illness (ILI, 3C), and respiratory syncytial virus (RSV, 3D), between 6 March 2017 and 13 March 2021.**

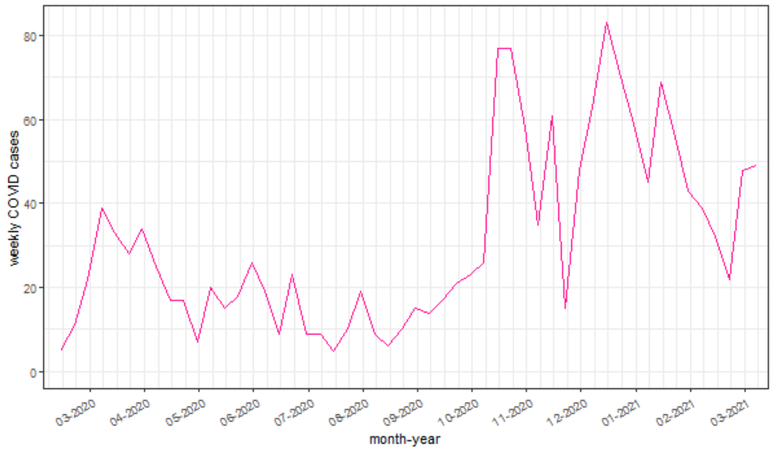


**Supplemental Figure 4. Weekly case count of COVID-19, between 6 March 2020 and including 13 March 2021.**

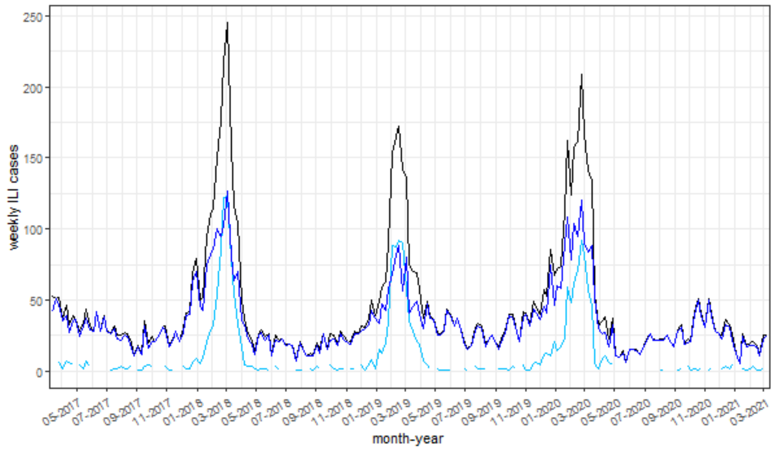


Supplemental Figure 5A: ILI cases (black), probable (dark blue), confirmed (light blue)

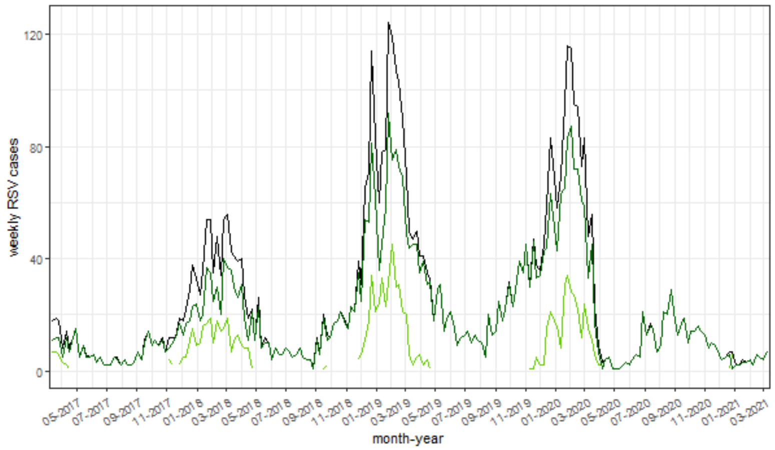


Supplemental Figure 5B: RSV cases (black), probable (dark green), confirmed (light green)

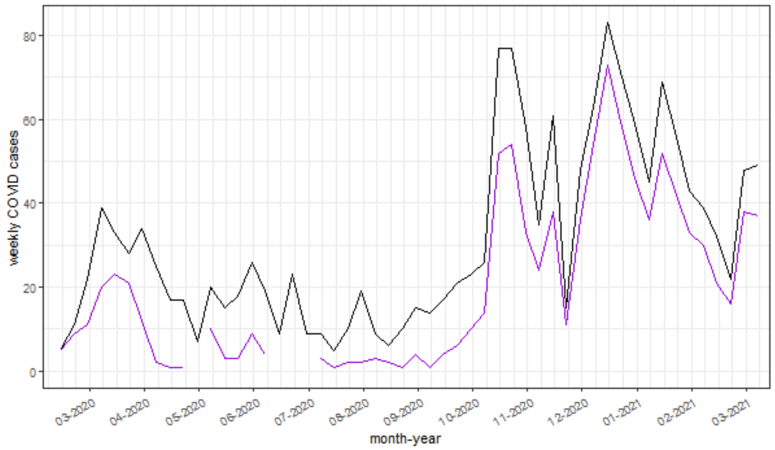


Supplemental Figure 5C: COVID-19 cases (black), confirmed (purple)

*The combined case definition of COVID-19 cases equals the probable case definition.*

**<u>Supplemental Figure 5.</u> Weekly case counts, by probable and possible case classifications as well as the combined case definition, of influenza-like-illness (ILI, 5A), respiratory syncytial virus (RSV, 5B), and COVID-19 (5C), between 6 March 2017 and 13 March 2021.**
